# Identifying Optimal COVID-19 Testing Strategies for Schools and Businesses: Balancing Testing Frequency, Individual Test Technology, and Cost

**DOI:** 10.1101/2020.10.11.20211011

**Authors:** Gregory D. Lyng, Natalie E. Sheils, Caleb J. Kennedy, Daniel Griffin, Ethan M. Berke

## Abstract

**Background:** COVID-19 test sensitivity and specificity have been widely examined and discussed yet optimal use of these tests will depend on the goals of testing, the population or setting, and the anticipated underlying disease prevalence. We model various combinations of key variables to identify and compare a range of effective and practical surveillance strategies for schools and businesses.

**Methods:** We coupled a simulated data set incorporating actual community prevalence and test performance characteristics to a susceptible, infectious, removed (SIR) compartmental model, modeling the impact of base and tunable variables including test sensitivity, testing frequency, results lag, sample pooling, disease prevalence, externally-acquired infections, and test cost on outcomes case reduction.

**Results:** Increasing testing frequency was associated with a non-linear positive effect on cases averted over 100 days. While precise reductions in cumulative number of infections depended on community disease prevalence, testing every 3 days versus every 14 days (even with a lower sensitivity test) reduces the disease burden substantially. Pooling provided cost savings and made a high-frequency approach practical; one high-performing strategy, testing every 3 days, yielded per person per day costs as low as $1.32.

**Conclusions:** A range of practically viable testing strategies emerged for schools and businesses. Key characteristics of these strategies include high frequency testing with a moderate or high sensitivity test and minimal results delay. Sample pooling allowed for operational efficiency and cost savings with minimal loss of model performance.

## INTRODUCTION

As schools and businesses re-open and attempt to stay open, promptly detecting people with infectious COVID-19 is essential, especially as the risk of transmission is expected to increase with colder weather, more time indoors, and closer contact with others.^1, 2^ Recommended actions to attenuate spread include symptom checking, monitoring underlying community prevalence, and responsive policy adjustment. In addition to robust public health measures, successful return to normalcy will be accelerated and hopefully sustained by optimal COVID-19 testing strategies. Despite being commonly recommended, little guidance suggests the right approach to testing and how best to balance cost, test selection, results delays, the value of sample pooling, and how changing local disease prevalence should inform strategy adjustments.

Throughout the pandemic the number and variety of tests for detecting active infection have steadily increased.^3^ Current tests include nucleic acid amplification tests (NAATs) such as reverse-transcription or reverse transcription polymerase chain reaction (RT-PCR), template mediated amplification (TMA), nicking enzyme amplification reaction (NEAR), loop-mediated isothermal amplification (LAMP), nucleic acid hybridization, viral metagenomic sequencing, and CRISPR-based assays. Most Food and Drug Administration (FDA) - Emergency Use Authorization (EUA) tests are approved for symptomatic patients, but not all are validated in an asymptomatic population. Despite these scientific advancements, there is scant guidance on how to apply a specific technology in the context of the underlying population and the goal of testing, such as diagnosis of an individual versus surveillance of a group. Cost, turnaround time, and convenience in sample collection all play a role in achieving a rate of testing that achieves a goal of detecting and preventing transmission in a cohort. A testing strategy is not feasible if the cost per test at the individual level is too high, or the time to obtain results is too long, resulting in possible transmission while positive test results are in transit or missing an opportunity to attend work or school if the result is negative. To increase test processing efficiency and reduce cost, pooling of samples is a potential solution provided there is minimal degradation in test performance due to dilution, but a strategy should be devised carefully. Successful pooling strategies rely on a clear understanding of the test’s limit of detection (LOD), sensitivity, specificity, and the prevalence of disease in the population being tested.^4^

Testing in large cohort settings such as schools and businesses that require continued surveillance can ensure that facilities remain open safely for the greatest number of people. We model various scenarios of test sensitivity and specificity, testing frequency, cost, and pooling to illustrate the range of practical and sustainable surveillance strategies.

## METHODS

To compare the effects of test sensitivity and specificity, test frequency, and the impact of pooling we considered a classical epidemiological susceptible, infectious, removed (SIR) compartmental model for the tested population. To account for the introduction of infections from the surrounding community, we added a time-dependent term which represents the rate (in people/time) of infections from outside interactions continuously in time. With frequent testing, this external forcing drives the behavior of the model (Figure 1). We examine two scenarios for this forcing. The first is a relatively low and more-or-less constant rate of introduced infections, with data from the 7-day rolling average of the case count in Fayette County, Pennsylvania for the 100 days beginning March 26, 2020 as reported in the New York Times.^5^ This low-growth profile is reported as panel (a) in Figures 2, 3, and 4. The second scenario used for high-growth external community prevalence is the seven-day rolling average of daily case counts in Miami-Dade County, Florida for the 100 days beginning June 16, 2020. This profile is shown in panel (b) in Figures 2, 3, and 4. In both profiles, we scaled the cases given by the relative population in our model, which we chose to be 1500. To model pooled testing we solved the SIR model over *τ* days, with the initial test on day zero. To account for possible delays in receiving test results, we allowed for a delay parameter, *d*. On day *τ* + *d* we stopped the model and restarted with new “initial conditions” which account for the transfer of people who tested positive and are thus removed from mixing in the model. We adjusted for test sensitivity and applied a linear discount rate for pooling of 0.00323, consistent with minimal sample dilution or degradation in a nasal or nasopharyngeal sample.^6^ Other discount rates may be more appropriate in different settings, such as saliva sampling.^7^ Our model allows for a varied percent of those that are infected to choose to comply with isolation protocols; in the scenarios presented we set this tunable assumption to be perfect compliance. We assume the basic reproduction number R_0_ is 2.5 and the average period of infectiousness is 9 days.^8, 9, 10, 11^ The initial conditions are chosen from the average of population-scaled new confirmed cases reported by the New York Times for September 23, 2020 in a sample of counties scaled by average number of infectious days. This results in a starting value of 1.35 infections for a population of size 1500. We take the conservative approach of assuming no one in the population has immunity to the virus based on previous infection. In the tests that follow we vary the testing frequency (*τ*), delay in the return of results (*d*), number of samples pooled (*m*), sensitivity of the test on one sample, and specificity of the test. We computed the cost of each testing strategy at the per person per day level, over 100 days. When pooling (*m* > 1), we assumed a simple 2-stage Dorfman testing process in which each individual in a positive pool is retested individually using a high-sensitivity diagnostic test at $100 per test. We then calculated the expected number of tests required to complete each round of testing. The complete scientific code is available as a supplementary file. All analysis was done using Julia v1.5.1.^12^

**Figure 1.**
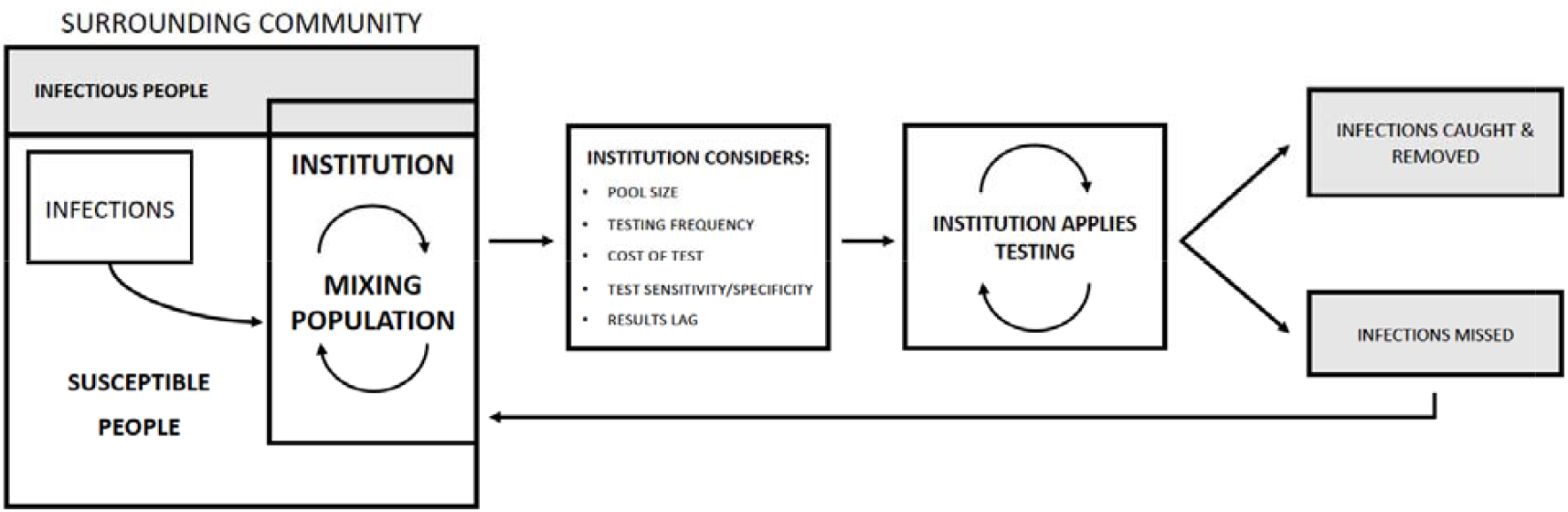
Schematic of the model. The model simulates testing for a common group of people who mix continuously in an institution (i.e., in a school or office) and are subject to the introduction of infection from the surrounding unmonitored community. The framework couples regular testing, described by a handful of tunable parameters, to a disease model. The disease model is dynamic in time, and infections may originate both from inside-the-institution mixing and from the surrounding community at varying rates depending on prevalence.

**Figure 2.**
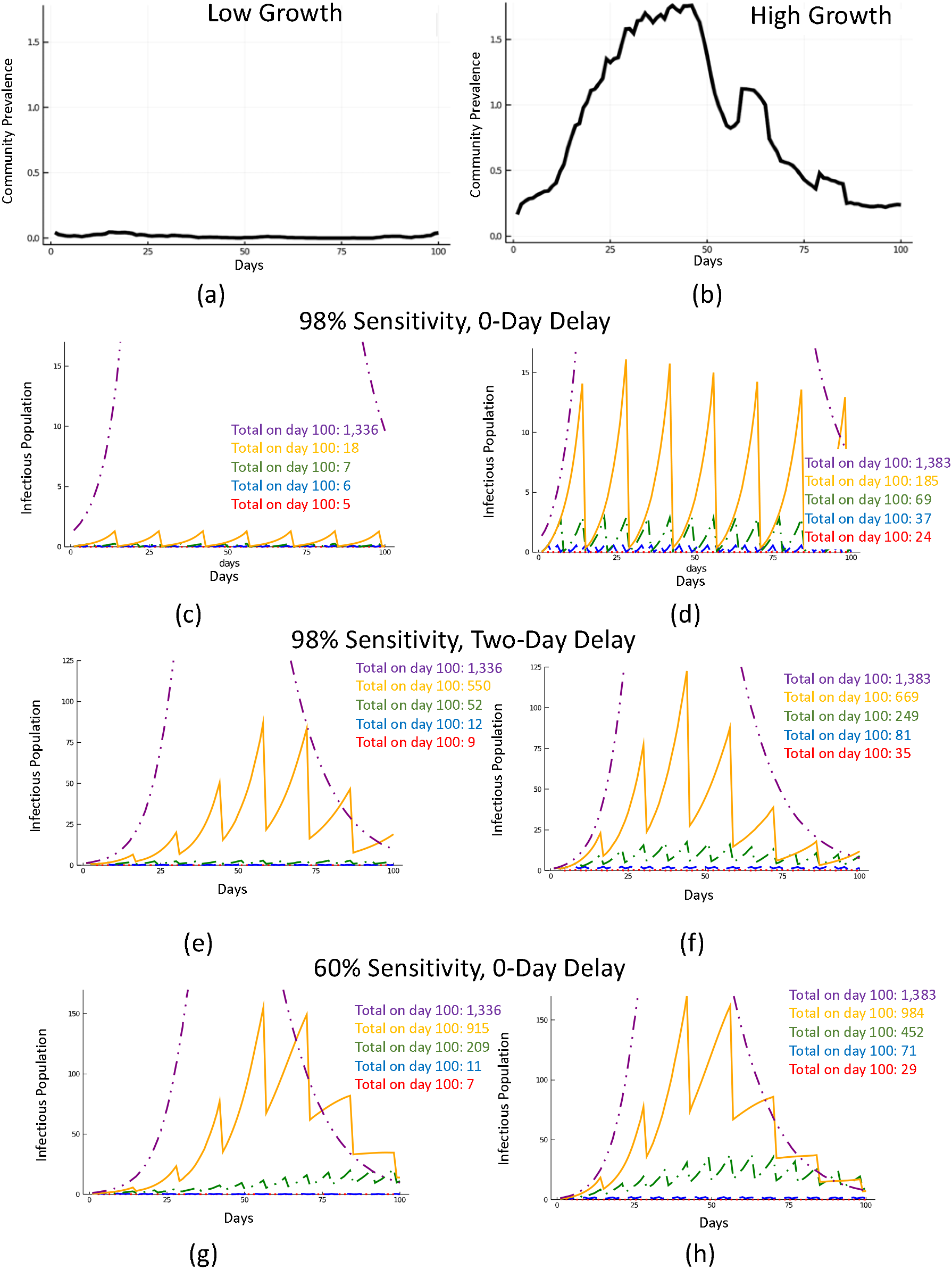
Impact of testing frequency. Two scenarios for community prevalence corresponding, relatively, to low and high rates of imported infections (Panels (a) and (b)). Testing with a test with 98% sensitivity with 0-day resulting delay amidst high and low community prevalence (Panels (c) and (d)). Testing with a test with 98% sensitivity with 2-day resulting delay amidst high and low community prevalence (Panels (e) and (f)). Testing with a test with 60% sensitivity with 0-day resulting delay amidst high and low community prevalence (Panels (g) and (h)). Uncropped figures are available in the supplement. Purple (dash-dot-dot) corresponds to no testing, orange (solid) to testing every two weeks, green for testing every week (dash-dot), blue (dash) for testing every 3 days, and red (dot) for daily testing.

**Figure 3.**
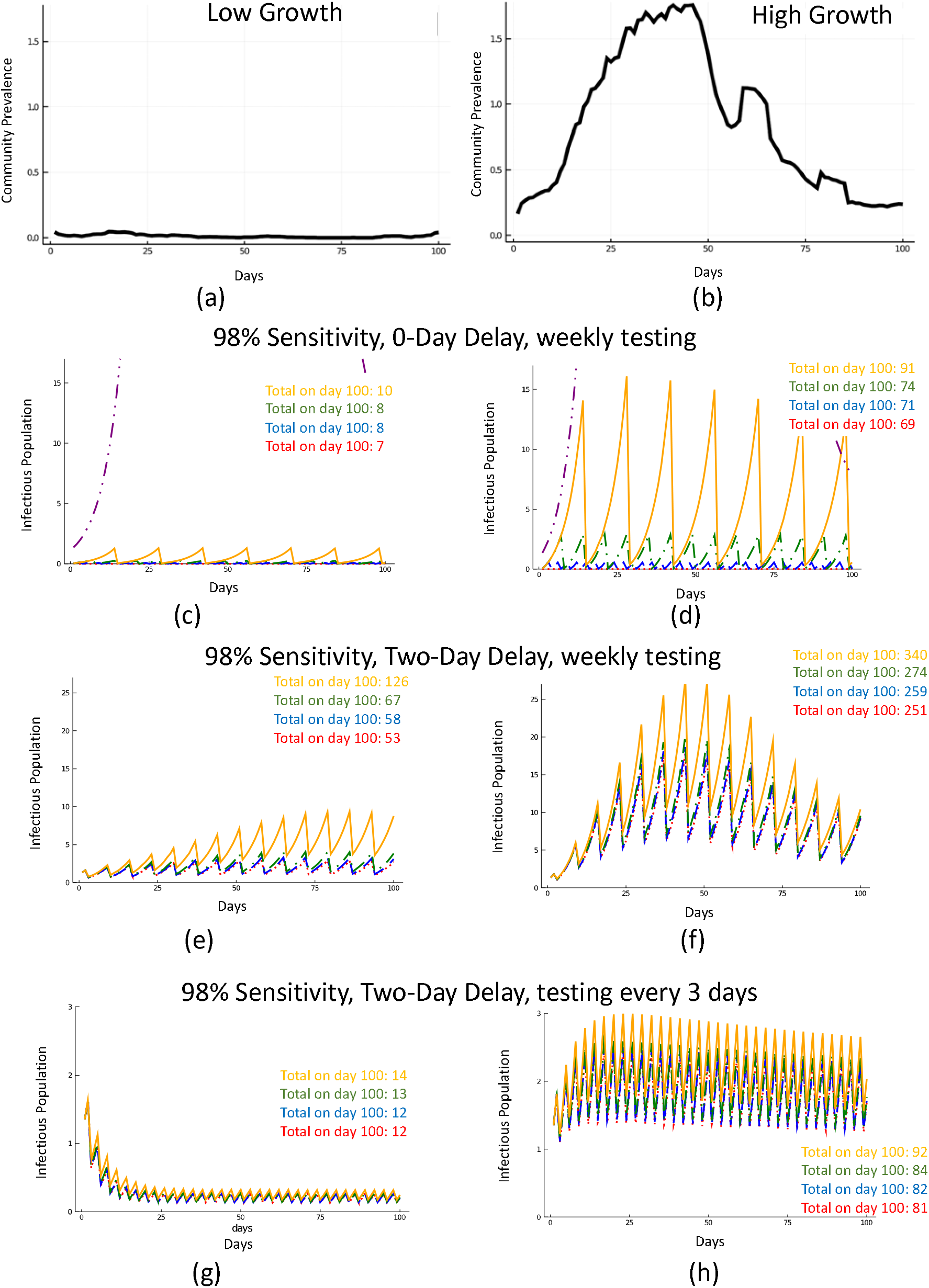
Effect of pool size. Two scenarios for community prevalence corresponding, relatively, to low and high rates of imported infections (Panels (a) and (b)). Testing weekly with a test with 98% sensitivity with 0-day resulting delay amidst high and low community prevalence (Panels (c) and (d)). Testing weekly with a test with 98% sensitivity with 2-day resulting delay amidst high and low community prevalence (Panels (e) and (f)). Testing ever 3 days with a test with 98% sensitivity with 2-day resulting delay amidst high and low community prevalence (Panels (g) and (h)). Orange lines (solid) correspond to 30 samples pooled, green (dash-dot) to ten samples pooled, blue (dash) to five samples pooled, and red (dot) to 2 samples pooled.

**Figure 4.**
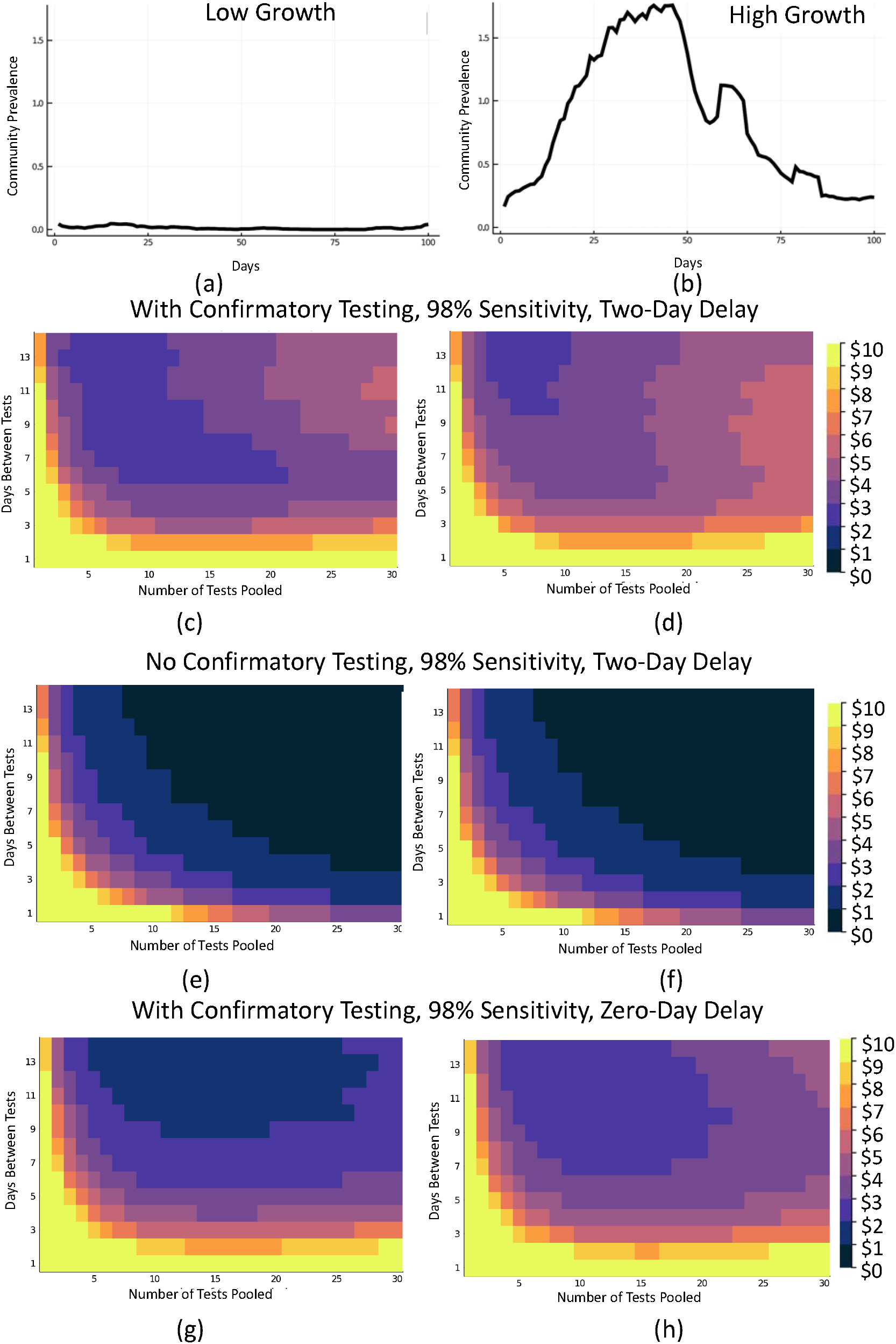
Cost comparison map for various pooling and frequency scenarios with and without confirmatory testing.* * Use case of a test with 98% sensitivity and 99.5% specificity with a 2-day result delay costing $100 and a 98% sensitive test with 99.5% specificity and a 0 day result delay costing $120. In (c,d, g, h) every person in a positive pool is retested for confirmation and in (e, f) no confirmatory testing is done. We assume all confirmatory tests cost $100. Colors correspond to cost per person per day in dollars.

## RESULTS

Figure 2 demonstrates scenarios of testing frequency at sensitivities of 98% with a two-day delay in receiving results during which mixing continues (Figure 2c, d), 98% with no delay in receiving results (Figure 2e, f), and 60% with no delay (Figure 2g, h) to simulate testing by various technologies such as PCR with lags between sample collection and centralized laboratory testing, antigen detection, and LAMP. As there are little data on the performance of some tests in asymptomatic people, we used more conservative sensitivity estimates aligning to published LOD for specific devices from the FDA.^13^ The sawtooth pattern is the result of removal of infected persons from the population.

Any testing strategy is better than none at all, and as expected, tests with increased sensitivities perform for a given time frequency. At the most lenient frequency considered, every 14 days, the number of infections is reduced approximately 31-98% (Table 1) compared to no testing at all. Each scenario can be explored comparatively. For example, at a test sensitivity of 80%, the effect of testing every day in a population of 1500 compared to testing every 14 days reduced the number of cumulative infections at day 100 by 364 in the low prevalence community and by 958 in the high prevalence community. Increased testing frequency results in a nonlinear decrease in cumulative infections over time, with daily testing resulting in the fewest cumulative infections at 100 days after implementing the testing strategy at any of the sensitivities shown. Importantly, at sensitivities of 98% our models predict that a two-day delay in results (by send-out PCR, for example) will result in a just a 59% reduction in infections experienced at a 14-day testing frequency; however, as the testing frequency is increased, even with the two-day delay, the number of missed infections goes down rapidly to over a 99% reduction from no testing at all at a daily testing frequency.

**TABLE 1.**
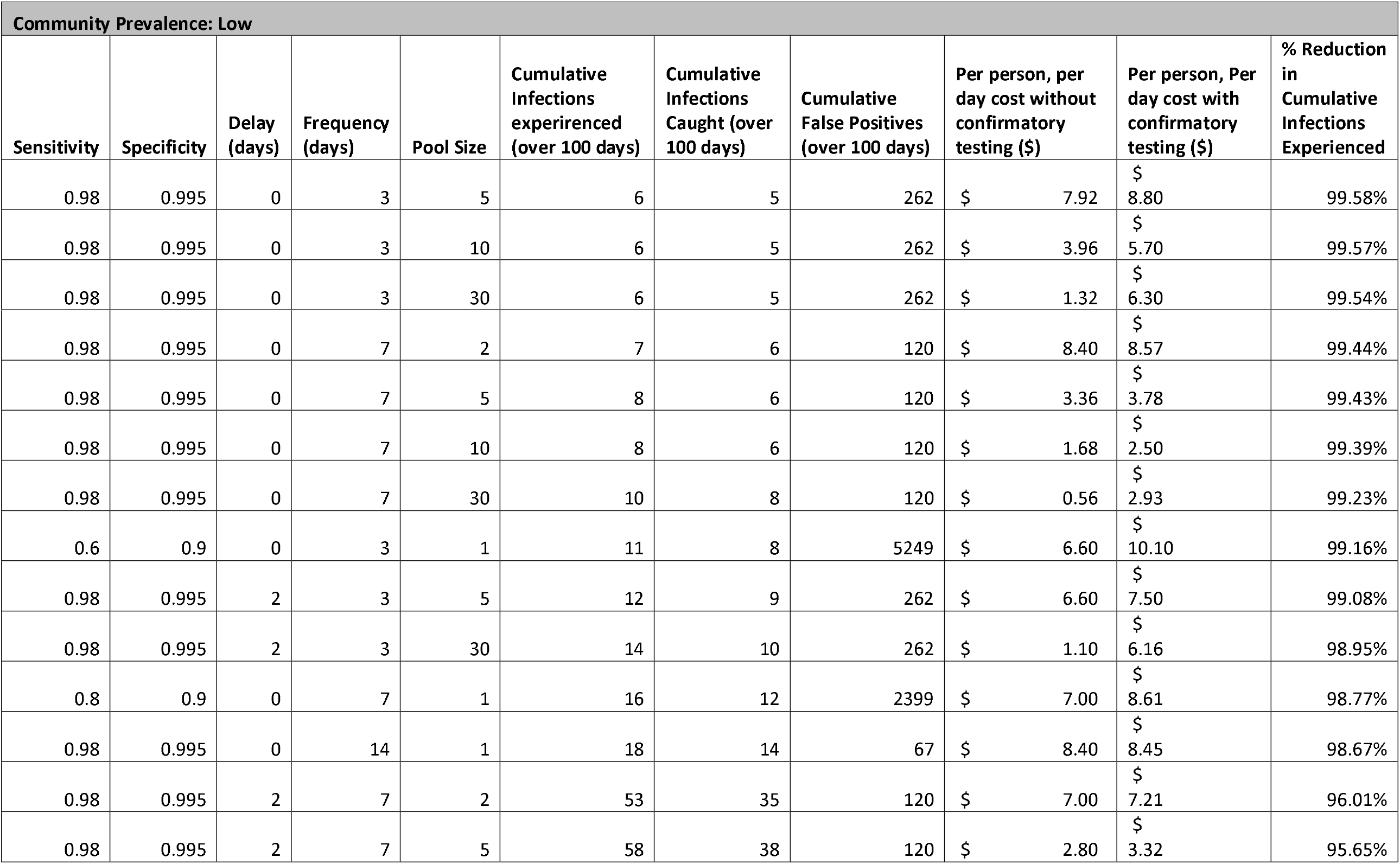
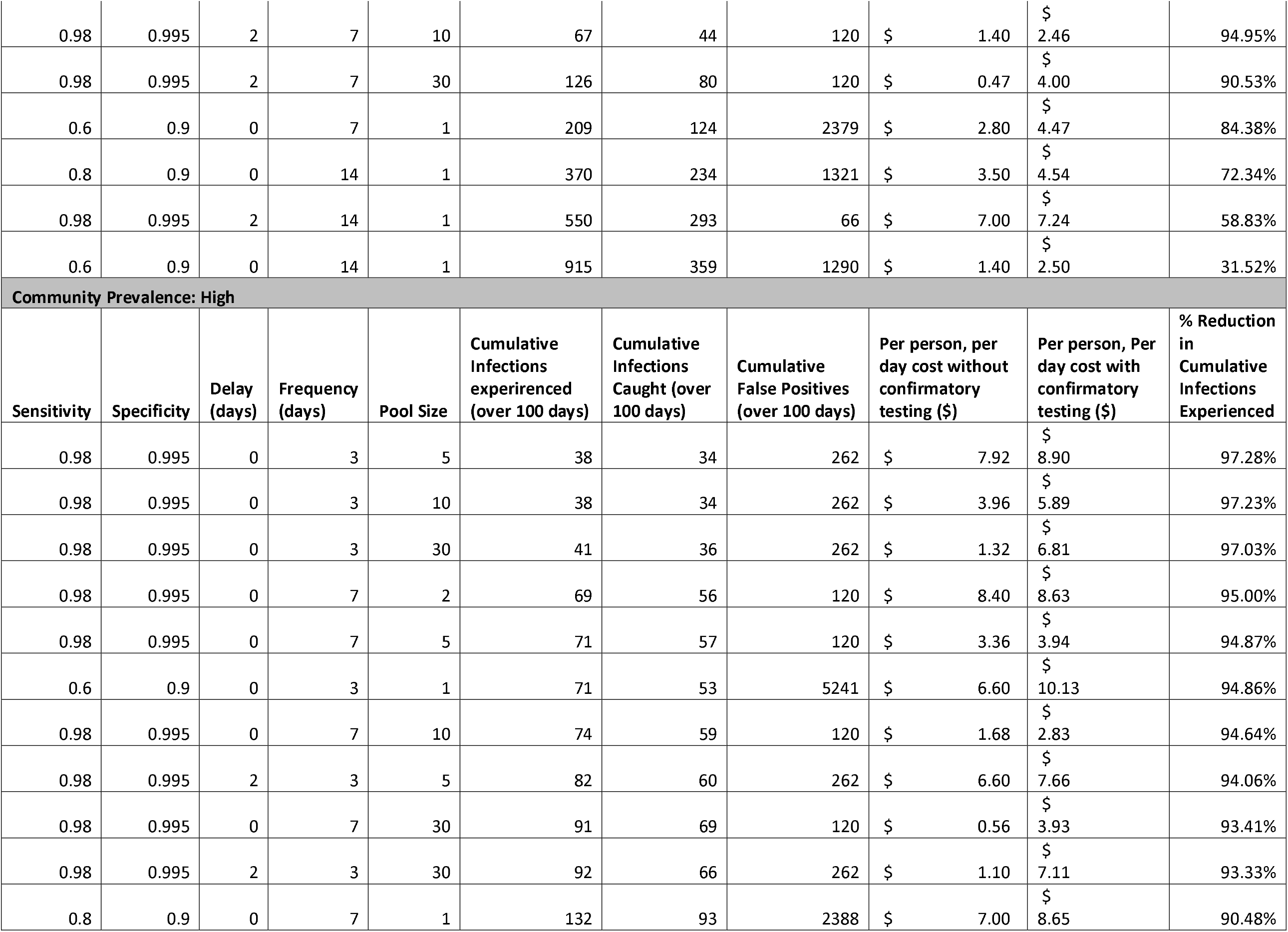
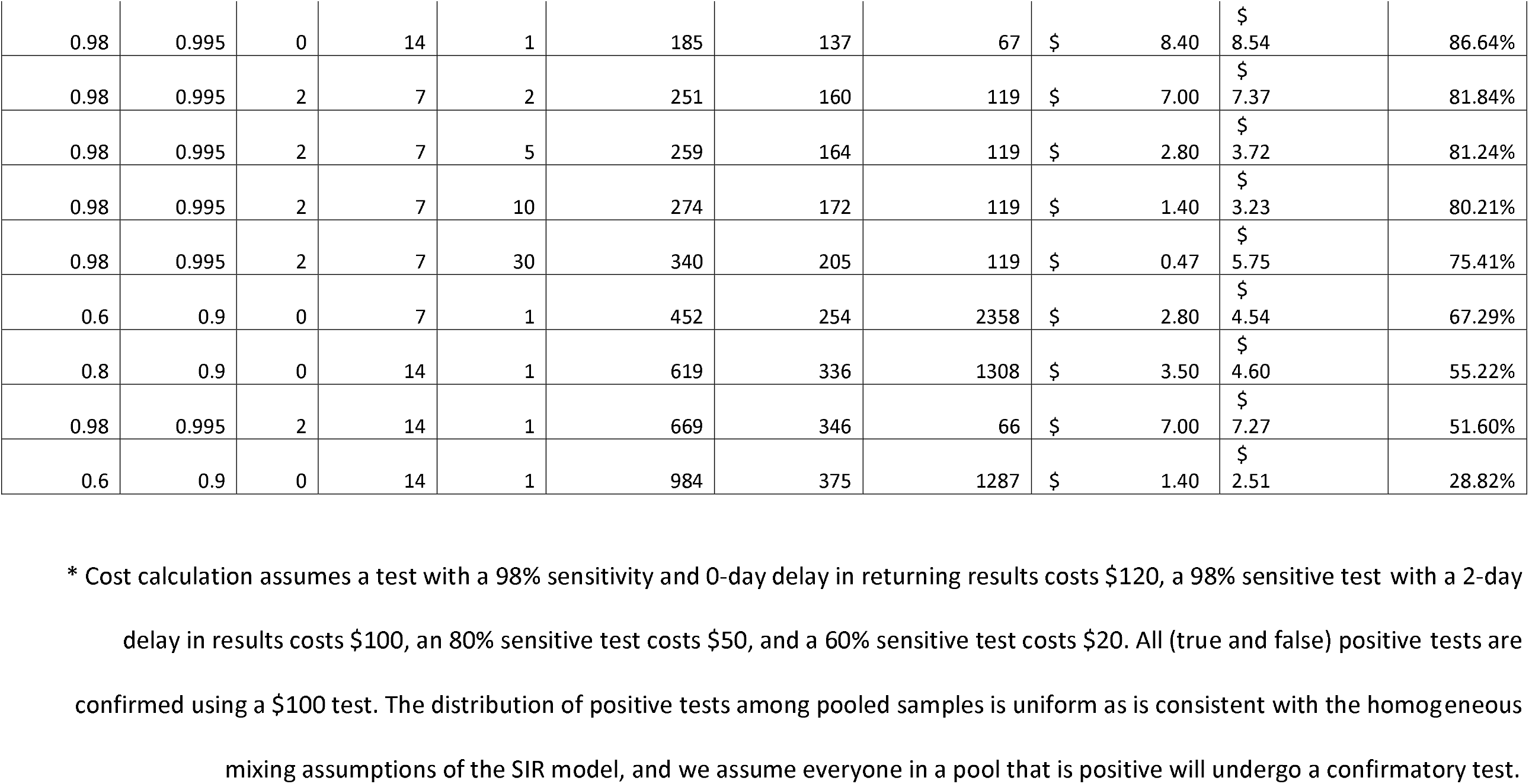
Selected testing strategies ranked by reduction in cumulative infections, with and without confirmatory testing, for scenarios costing less than $10 per person per day. *

Next we looked at testing strategies that incorporate pooling. Figure 3 combines a weekly and every 3 day testing strategy with 98% sensitive tests with varying time delay, and pooling tests in samples of 2, 5, 10, and 30. Pooling potentially reduces the sensitivity of the tests, resulting in more missed infections. This can be overcome by an increase in test frequency, allowed by the cost savings of pooling. Figure 4 weighs cost against testing frequency and pooling size, both with confirmatory and without confirmatory testing of positive pools. Without confirmatory testing, the cost per person decreases dramatically.

## DISCUSSION

Our findings demonstrate that it is not only critical to choose the right test in terms of performance in asymptomatic individuals, but to use the test in the defined population at the optimal frequency to reduce the risk of case escalation. Optimization is further enhanced at the population level by understanding of underlying disease prevalence and utilization of pooling to reduce cost and increase efficiency. The “ideal” test strategy must be balanced with the practicalities of cost per person to ensure sustainability. For example, daily testing with a 60% sensitive test attenuates community spread, but at a cost of $30.10 per person per day with confirmatory testing, or $20.00 without, may not be possible. Using a 60% sensitive test less frequently reduces expense but sacrifices significant performance. A 98% sensitive test with no delay in results administered every 3 days with pooling, and no confirmatory test offered by the institution costs less than $1.50 per person per day, with high performance. The model demonstrates that frequency of testing, test sensitivity, turn-around time, and the external community prevalence are all important factors to consider, and there is often more than one testing strategy to achieve the desired level of performance. The computational code and an is available in the online supplement and an easy-to-use web-based simulator is to test various scenarios at https://calculator.unitedinresearch.com/.

With these scenarios in hand, institutions can make an informed operational choice, devise pods or cohorts to be tested by pooling and potentially isolated if positive, and create clear communication about a surveillance rationale. Acknowledging a dynamic community prevalence, the model can be re-run, and the testing strategy can be optimized to maximize benefit at the lost cost and least amount of disruption.

The frequency of test usage to minimize amplification of infection and allow schools and worksites to remain open is an important factor. Given the cost of high frequency testing, we demonstrate the value of pooling of samples to increase efficiency, particularly in areas with lower population prevalence. As background prevalence increases, the value of pooling diminishes as the likelihood of a positive pool will rise, but even a pool of two to three samples results in a dramatic reduction in the need for individual sample analysis. It is worth noting that with an extremely low prevalence, even in the case of a 99.5% specific test, false positives are much more likely than true positives and confirmatory testing may be necessary. A 90% specificity test would result in a large number of false positives over the course of 100 days. As shown in Figure 4, in order to achieve a minimal cost approach that includes confirmatory testing, one must balance pool size with frequency. Without confirmatory testing, costs drop dramatically (Figure 4g, h). More sophisticated confirmatory testing strategies exist that may lower costs but still reduce the likelihood that uninfected individuals are sent home, such as sub-pooling of positive pools without individual level testing, each with benefits and disadvantages.^14, 15^

Our work is supported by prior discoveries. Paltiel et al.^16^ considered a compartment-based model simulating an abbreviated 80-day semester in a highly-residential college-campus-type setting. Across all scenarios considered, test frequency was more associated with cumulative infection than test sensitivity. That modeling exercise also suggested that symptom-based screening alone is insufficient to contain an outbreak under any of the scenarios considered. Using a model for viral loads in individuals, Larremore et al.^17^ studied surveillance effectiveness using an agent-based modeling framework which accounts for test sensitivities, frequency, and sample-to-answer reporting time. The results indicate that frequency of testing and the speed of reporting are the principal contributors to surveillance effectiveness. The results also show that the impact of high sensitivity on surveillance effectiveness is, relatively, small.

Populations housed in long-term care facilities are especially vulnerable to COVID-19; surveillance programs designed for these settings may have different goals and tolerances for infection risk than those designed to maintain functionality for other institutions. Smith and colleagues^6^ built a complex modeling framework for long-term care facilities including simulations of the detailed inter-individual contact networks describing patient-staff interactions in such settings. This work showed that symptom-based screening by itself had limited effectiveness. Testing upon admission detected most asymptomatic cases upon entry but missed potential introductions from staff. Random daily testing was determined to be, overall, an inefficient use of resources. This points to the opportunity for pooled testing as an effective and efficient COVID-19 surveillance strategy for long-term care facilities with limited resources.

Since our work focuses on screening and not performing diagnostic testing, the actual sensitivity of the various available COVID tests for this purpose is not entirely clear. The original testing approaches for COVID-19 focused on the high sensitivity required for diagnosis by clinicians in all stages of the acute period of COVID-19 through detection of SARS-CoV-2 RNA performed on patients with a high pretest probability of disease. This paradigm focused on high sensitivity tests with the performance feature of very low NAAT detectable units/mL (NDU/mL) with a goal of diagnosing patients even if past the contagious period. These tests were not optimized nor validated in terms of sensitivity for the detection of infectious individuals that might spread disease in schools, the workplace or other social situations.

Several studies looking at the ability to culture virus from samples collected from infected individuals have established that RNA copy numbers of 1,000,000 RNA copies/ml or higher are required for any consistent success in viral culture.^18, 19, 20, 21, 22^ Based on contact tracing, this defined window of elevated RNA copy numbers starting 2-3 days prior to onset of symptoms and ending 5-9 days after symptom onset corresponds to most if not all cases of transmission. Studies of asymptomatic spreading suggests a very similar window of transmissibility during this period of time when RNA copy numbers are 1,000,000 copies/ml or higher.^11, 23, 24^ Given that RT-PCR testing can have a sensitivity or LOD as low as <1,000 RNA copies/mL (1,000 NDU), there should ample performance in testing technology to leverage high-volume, high-frequency pooling, provided samples are not diluted by storage or buffering media beyond the minimum LOD when employed to detected asymptomatic but infectious individuals ^25^

Our work has a number of limitations. The SIR compartmental model provides a simplified representation of the natural history of the disease. For example, it does not account for the distinction between symptomatic and asymptomatic cases. In addition, the model assumes uniform mixing of the population being tested and a uniform distribution of likelihood of a positive test. The model is formulated at a population level; it does not permit the tracking of individuals. In a low population prevalence, we expect a high number of false positives given assumed specificities of 99.5% and 90%. Individuals who recover from the disease are granted permanent immunity in our model, although the risk of reinfection now appears possible.^26, 27, 28, 29, 30, 31^ Our pooling model assumed nasal or naso-pharyngeal swab samples. Because of the nature of saliva, the small sensitivity discount rate assumption in our model may not be valid due to greater sample dilution.^32^ Finally, the model does not naturally incorporate phased, pulsed, or partial testing (1^st^ graders on Monday, 2^nd^ graders on Tuesday, etc.).

Despite these limitations, sensitivity, pooling, and frequency modeling can guide institutions on best-fit testing strategies that align to their practical constraints. Organizations can apply this model to determine their best testing strategy given current community prevalence and operational and financial resources that enable sustained testing to stay safely open during the pandemic.

## Data Availability

Computational code is included in the on-line supplement.

https://calculator.unitedinresearch.com/

## REFERENCES

1. Service RF. Coronavirus antigen tests: quick and cheap, but too often wrong? Science | AAAS. Published May 22, 2020. Accessed September 23, 2020. https://www.sciencemag.org/news/2020/05/coronavirus-antigen-tests-quick-and-cheap-too-often-wrong

2. Corman VM, Rabenau HF, Adams O, et al. SARS-CoV-2 asymptomatic and symptomatic patients and risk for transfusion transmission. Transfusion. 2020;60(6):1119–1122. doi:10.1111/trf.15841

3. FDA. Emergency Use Authorization. FDA. Published online September 22, 2020. Accessed September 23,2020. https://www.fda.gov/emergency-preparedness-and-response/mcm-legal-regulatory-and-policy-framework/emergency-use-authorization

4. Abdalhamid B, Bilder CR, McCutchen EL, Hinrichs SH, Koepsell SA, Iwen PC. Assessment of Specimen Pooling to Conserve SARS CoV-2 Testing Resources. Am J Clin Pathol. Published online April 18, 2020. doi:10.1093/ajcp/aqaa064

5. The New York Times. NYT COVID-19 Data. COVID-19 Data. Published September 23, 2020. Accessed September 23, 2020. https://github.com/nytimes/covid-19-data

6. Smith, David RM, Duval, Audrey, Pouwels, Koen B, et al. Optimizing COVID-19 surveillance in long-term care facilities: a modelling study. Published online August 13, 2020. https://www.medrxiv.org/content/10.1101/2020.04.19.20071639v2

7. Watkins AE, Fenichel EP, Weinberger DM, et al. Pooling Saliva to Increase SARS-CoV-2 Testing Capacity. Infectious Diseases (except HIV/AIDS); 2020. doi:10.1101/2020.09.02.20183830

8. Wu JT, Leung K, Leung GM. Nowcasting and forecasting the potential domestic and international spread of the 2019-nCoV outbreak originating in Wuhan, China: a modelling study. The Lancet. 2020;395(10225):689–697. doi:10.1016/S0140-6736(20)30260-9

9. He X, Lau EHY, Wu P, et al. Temporal dynamics in viral shedding and transmissibility of COVID-19. Nature Medicine. 2020;26(5):672–675. doi:10.1038/s41591-020-0869-5

10. Griffin JM, Collins AB, Hunt K, et al. A rapid review of available evidence on the serial interval and generation time of COVID-19. medRxiv. Published online January 1, 2020:2020.05.08.20095075. doi:10.1101/2020.05.08.20095075

11. Ferretti L, Ledda A, Wymant C, et al. The timing of COVID-19 transmission. medRxiv. Published online September 16, 2020:2020.09.04.20188516. doi:10.1101/2020.09.04.20188516

12. Bezanson J, Edelman A, Karpinski S, Shah VB. Julia: A Fresh Approach to Numerical Computing. SIAM Rev. 2017;59(1):65–98. doi:10.1137/141000671

13. Health C for D and R. SARS-CoV-2 Reference Panel Comparative Data. FDA. Published online September 15, 2020. Accessed September 25, 2020. https://www.fda.gov/medical-devices/coronavirus-covid-19-and-medical-devices/sars-cov-2-reference-panel-comparative-data

14. Bilder CR. Group Testing for Identification. In: Wiley StatsRef: Statistics Reference Online. American Cancer Society; 2019:1–11. doi:10.1002/9781118445112.stat08227

15. Chan CL, Jaggi S, Saligrama V, Agnihotri S. Non-Adaptive Group Testing: Explicit Bounds and Novel Algorithms. IEEE Transactions on Information Theory. 2014;60(5):3019–3035. doi:10.1109/TIT.2014.2310477

16. Paltiel AD, Zheng A, Walensky RP. Assessment of SARS-CoV-2 Screening Strategies to Permit the Safe Reopening of College Campuses in the United States. JAMA Netw Open. 2020;3(7):e2016818–e2016818. doi:10.1001/jamanetworkopen.2020.16818

17. Larremore DB, Wilder, Bryan, Lester, Evan, et al. Test sensitivity is secondary to frequency and turnaround time for COVID-19 surveillance. https://www.medrxiv.org/content/10.1101/2020.06.22.20136309v3

18. Bullard J, Dust K, Funk D, et al. Predicting Infectious Severe Acute Respiratory Syndrome Coronavirus 2 From Diagnostic Samples. Clin Infect Dis. doi:10.1093/cid/ciaa638

19. He D, Zhao S, Lin Q, et al. The relative transmissibility of asymptomatic COVID-19 infections among close contacts. International Journal of Infectious Diseases. 2020;94:145–147. doi:10.1016/j.ijid.2020.04.034

20. Huang C-G, Lee K-M, Hsiao M-J, et al. Culture-Based Virus Isolation To Evaluate Potential Infectivity of Clinical Specimens Tested for COVID-19. Journal of Clinical Microbiology. 2020;58(8). doi:10.1128/JCM.01068-20

21. Perera RAPM, Tso E, Tsang OTY, et al. SARS-CoV-2 Virus Culture and Subgenomic RNA for Respiratory Specimens from Patients with Mild Coronavirus Disease. Emerging Infectious Diseases. 26(11). doi:10.3201/eid2611.203219

22. Singanayagam A, Patel M, Charlett A, et al. Duration of infectiousness and correlation with RT-PCR cycle threshold values in cases of COVID-19, England, January to May 2020. Eurosurveillance. 2020;25(32):2001483. doi:10.2807/1560-7917.ES.2020.25.32.2001483

23. Mizumoto K, Kagaya K, Zarebski A, Chowell G. Estimating the asymptomatic proportion of coronavirus disease 2019 (COVID-19) cases on board the Diamond Princess cruise ship, Yokohama, Japan, 2020. Eurosurveillance. 2020;25(10):2000180. doi:10.2807/1560-7917.ES.2020.25.10.2000180

24. Savvides, Christina, Siegel, Robert. Asymptomatic and presymptomatic transmission of SARS-CoV-2: A systematic review. 2:https://www.medrxiv.org/content/10.1101/2020.06.11.20129072v2

25. Zhen W, Manji R, Smith E, Berry GJ. Comparison of Four Molecular In Vitro Diagnostic Assays for the Detection of SARS-CoV-2 in Nasopharyngeal Specimens. Journal of Clinical Microbiology. 2020;58(8). doi:10.1128/JCM.00743-20

26. Kirkcaldy RD, King BA, Brooks JT. COVID-19 and Postinfection Immunity: Limited Evidence, Many Remaining Questions. JAMA. 2020;323(22):2245–2246. doi:10.1001/jama.2020.7869

27. Xing Y, Mo P, Xiao Y, Zhao O, Zhang Y, Wang F. Post-discharge surveillance and positive virus detection in two medical staff recovered from coronavirus disease 2019 (COVID-19), China, January to February 2020. Eurosurveillance. 2020;25(10):2000191. doi:10.2807/1560-7917.ES.2020.25.10.2000191

28. Bao L, Deng W, Gao H, et al. Lack of Reinfection in Rhesus Macaques Infected with SARS-CoV-2. bioRxiv. Published online May 1, 2020:2020.03.13.990226. doi:10.1101/2020.03.13.990226

29. Xiao AT, Tong YX, Zhang S. False negative of RT-PCR and prolonged nucleic acid conversion in COVID-19: Rather than recurrence. Journal of Medical Virology. 2020;92(10):1755–1756. doi:10.1002/jmv.25855

30. Lan L, Xu D, Ye G, et al. Positive RT-PCR Test Results in Patients Recovered From COVID-19. JAMA. 2020;323(15):1502-1503. doi:10.1001/jama.2020.2783

31. Wu J, Liu X, Liu J, et al. Coronavirus Disease 2019 Test Results After Clinical Recovery and Hospital Discharge Among Patients in China. JAMA network open. 2020;3(5):e209759. doi:10.1001/jamanetworkopen.2020.9759

32. Watkins AE, Fenichel EP, Weinberger DM, et al. Pooling saliva to increase SARS-CoV-2 testing capacity. medRxiv. Published online September 3, 2020:2020.09.02.20183830. doi:10.1101/2020.09.02.20183830

