## Supplemental Methods & Code for "Identifying Optimal COVID-19 Testing Strategies for Schools and Businesses: Balancing Testing Frequency, Individual Test Technology, and Cost"

### 1. Overview

We give a detailed description of the model underpinning the analysis for testing schools and businesses. The modeling framework describes a scenario in which there is monitoring/testing for a common group of people who mix continuously (as in a school or office setting), and are subject to the introduction of infection from the surrounding (unmonitored) community. All together, the model links a testing strategy, described by a number of tunable parameters, to a disease model for COVID-19. The disease model is dynamic in time as infections spread both from internal mixing and from the surrounding community. The tunable testing parameters correspond to attributes of the tests themselves (like sensitivity and specificity) and also to the elements of the strategy (like how many days between tests). By running all combinations of these parameters and counting the expected number of tests required, we may estimate the costs of each distinct surveillance strategy. We may also compare the strategy to the disease model running without implementing any testing at all to measure the effectiveness of the strategy. We begin by describing the testing strategies considered, and then describe the disease model in detail.

### 2. Testing strategies

2.1. **Test attributes and tunable parameters.** Our first assumption is that testing will happen on a regular cadence. Every day. Every 2 days. Weekly. Moreover, we assume that every individual in the organization is tested during each round of testing. Each test is characterized by four numbers:

- Sensitivity (Se) and Specificity (Sp),
- Cost ($C$) in dollars/test, and
- results lag $d$ in days.

Point-of-care tests feature $d = 0$, while, in the manuscript, traditional lab-based tests are assumed to have $d = 2$. Other values of $d$ may be appropriate to local circumstances. The two main choices of the organization are the frequency of testing (how many days between tests, $\tau - 1$) and the number of samples to pool ($m$, if pooling is an option for that particular test). Finally, the model allows less than 100% of individuals with positive tests to comply with isolation protocols. While this number ($b$) is not determined by organizational leaders, they will be in the best position to estimate realistic values for compliance in their organization. Another decision to be made at an organizational level concerns confirmatory testing. In low prevalence scenarios, especially with a non-specific test, the number of false positives will be large. An organizational commitment to fund confirmatory testing may involve substantial expense.

Tunable testing parameters for the model are listed in Table 1.

2.2. **Pooling.** Our quantitative approach to pooling directly follows the work of Smith et al. [3]. In that work (and in ours), we imagine samples (swabs) from multiple people being pooled and tested all at once. In low prevalence settings, this kind of group testing is widely used in human infectious disease applications, and the simple version we propose here was used by the US military to screen new inductees for syphilis during WWII [1]. In many circumstances, it is possible to greatly reduce

Table 1. Tunable testing parameters

| Parameter | Label | Description |
|---|---|---|
| Test frequency | $\tau$ | $\tau - 1$ days between tests |
| Results delay | $d$ | no. of days to wait for results |
| Samples pooled | $m$ | no. of samples in a pool |
| Compliance | $b$ | fraction of individuals who isolate after a positive test |
| Sensitivity | Se | individual-level sensitivity of the test |
| Specificity | Sp | individual-level sensitivity of the test |
| Cost | $C$ | cost of single test (in US dollars) |

the number of tests required to screen a population. However, there is a cost; pooled testing will result in decreased sensitivity.

Yelin et al. [4], using standard RT-PCR, estimated a sensitivity of

$$\mathrm{Se} = 0.9$$

after pooling a single positive SARS-CoV-2-positive sample together with 31 negative samples. We assume that there are no false negatives in the collection of un-pooled samples. We further assume a linear decrease in sensitivity with respect to negative samples added to the pool. Then, it is a simple matter to calculate the discount rate in sensitivity per each additional negative sample:

$$r = \frac{s_0 - s_{31}}{31} \,. \tag{2.1}$$

In equation (2.1) $s_0$ is the test sensitivity when no true negative samples were pooled with the original true positive sample, $s_{31}$ is the sensitivity when 31 true negative samples were pooled with the positive sample. Arithmetic yields $r = .00323$. Finally, for a pooled test with $s$ samples and $p$ true positive samples, the sensitivity can be calculated as

$$\mathrm{Se}_{\mathrm{gp}} = \mathrm{Se}_{\mathrm{ind}} - r(s - p) \,. \tag{2.2}$$

Here, $\mathrm{Se}_{\mathrm{gp}}$ represents the sensitivity of the pooled test and $\mathrm{Se}_{\mathrm{ind}}$ is the sensitivity of the individual-level test. In practice, of course, we don't know the number of true positives in the pool, so we assume the same prevalence of infection as in the monitored population. Individual RT-PCR testing for SARS-CoV-2 is highly specific, so a specificity of $\mathrm{Sp} = 0.995$ was assumed for all PCR-type tests regardless of the number of samples pooled.

When pooling ($m > 1$) and considering confirmatory testing, we assumed a simple 2-stage Dorfman testing process in which each individual in a positive pool is retested individually using a high-sensitivity diagnostic test. We then calculated the expected number of tests required to complete each round of testing. That is, suppose $X$ is the random variable counting the number of tests required to complete 2-stage pooling. Then, evidently, the quantity of interest is $\mathbb{E}[X]$, the expected number of tests need to complete one round of Dorfman 2-stage pooled testing. This will depend on the prevalence $p$ and on the sensitivity Se and specificity Sp.

For an individual test, the probability of returning a positive result is

$$q = \mathrm{Se}p + (1 - \mathrm{Sp})(1 - p) \,,$$

and the probability of returning a negative result is

$$1 - q = (1 - \mathrm{Se})p + \mathrm{Sp}(1 - p) \,.$$

We treat each test in the pool as an independent Bernoulli trial with probability of success $q$, and we denote by $Y$ the random variable counting the number of successful trials in a pool of $m$ samples.

By elementary probability,

$$\mathbb{P}(Y = k) = \binom{m}{k} q^k (1-q)^{m-k}, \tag{2.3}$$

whence $\pi_m$, the probability of having at least one positive test in the pool, is given by

$$\pi_m = \mathbb{P}(Y \geq 1) = 1 - \mathbb{P}(Y = 0) = 1 - [\mathrm{Sp} + p(1 - \mathrm{Se} - \mathrm{Sp})]^m . \tag{2.4}$$

Finally, if a pool tests positive, then $m$ additional confirmatory tests are required. Otherwise, only the single pool-level test is needed. Therefore,

$$\mathbb{E}[X] = \frac{P}{m}\left[m\pi_m + 1\right] . \tag{2.5}$$

### 3. Disease Model

3.1. **Population characteristics and disease.** We assume a monitored population of $P$ individuals. To characterize the disease, we use a classical continuous-time SIR model from classical epidemiology. That is, the monitored population is divided into compartments

- $S$ — susceptible,
- $I$ — infectious,
- $R$ — removed.

The disease model is characterized by two parameters: $R_0$ and $\gamma$. The basic reproduction number is well known, and we assume $R_0 = 2.5$ in agreement with early estimates of the reproduction number for COVID-19. The second parameter, $\gamma$, defines the removal rate for the disease, and $\gamma^{-1}$ is the average period of infectiousness. We assume $\gamma^{-1} = 9$ days.

Because the disease model is formulated as a system of differential equations, we must supply initial conditions, namely the values of $S$, $I$, and $R$ at the initial time $t_0$. In the manuscript, the initial conditions are chosen from the average of population scaled new confirmed cases reported by the New York Times for September 23, 2020 in a sample of counties scaled by $1/\gamma$. That is, we begin with 1.35 infections on day 0. We take the conservative approach of assuming no one in the population has immunity to the virus based on previous infection or otherwise. Thus $R(t_0) = 0$.

3.2. **Equations.** The core of the model is formed by the simple nonlinear, nonautonomous system of ordinary differential equations. (Note $\beta/\gamma = R_0$.)

$$\frac{\mathrm{d}}{\mathrm{d}t} S = -\beta S I - \epsilon(t) , \tag{3.1a}$$

$$\frac{\mathrm{d}}{\mathrm{d}t} I = \beta S I - \gamma I + \epsilon(t) , \tag{3.1b}$$

$$\frac{\mathrm{d}}{\mathrm{d}t} R = \gamma I . \tag{3.1c}$$

In (3.1), the forcing term $\epsilon$ accounts for the introduction of infections from outside the organization; see §3.5 for and explanation and derivation.

3.3. **Implementation.** To model pooled testing we solved the SIR model over $\tau$ days, with the initial test on day zero. To account for possible delays in receiving test results, we allow for a delay parameter, $d$. On day $\tau + d$ we stopped the model and restarted with new "initial conditions" which account for the transfer of the number of people who tested positive from the infectious to the removed compartment. This process is repeated according to the testing strategy defined by $\tau$, $m$, $d$, Sp, Se, and the total number of tests administered and infections caught are recorded. Rounding happens at each stoppage to estimate the number of positive pools and tests taken and, at the end of the simulation, to return whole number values for the number of infections caught.

3.4. **The cost of doing nothing.** To measure the effectiveness of the various strategies, we can compare them to the strategy of doing no testing. This gives one easily quantified measure, in terms of the reduction in cumulative infections over the time period, for evaluating the performance of a strategy. Figure 1 illustrates the dynamics of the model in the absence of testing. Likewise, Figure 2 reproduces Figure 2 from the main manuscript without any cropping; it illustrates the magnitude of the difference between an outbreak mitigated by testing and removal versus one checked by any kind of testing and isolation regime.

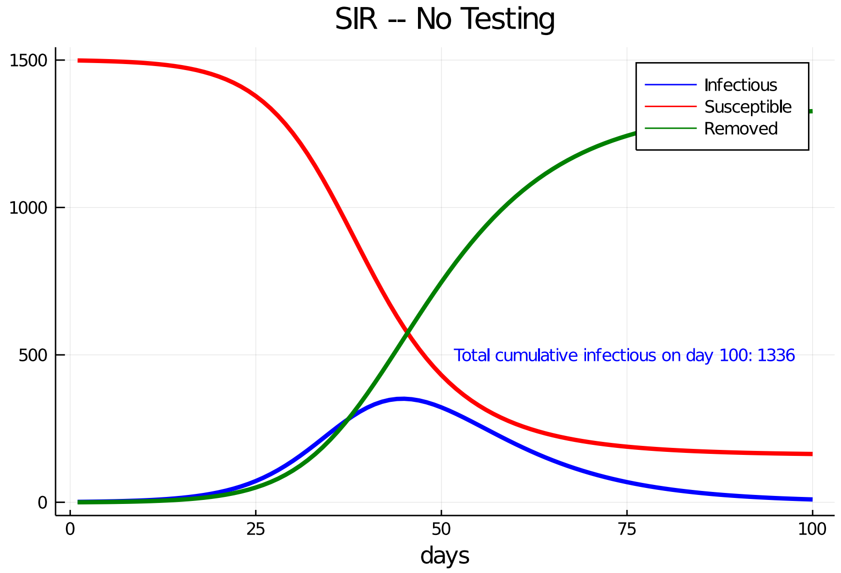


FIGURE 1. The cost of doing nothing. Running the forced model without testing, in this case with the flat profile, illustrates SIR-type dynamics with infections touching virtually the entire population.

3.5. **Community prevalence & time-dependent forcing.** To account for the introduction of infections from the surrounding community, we add a time-dependent forcing term which represents the rate of people becoming infected from an external source continuously in time. With frequent testing, this external forcing drives the behavior of the model. In general, the forcing takes the form of function

$$\epsilon : [0, \infty) \to \mathbb{R}\,, \tag{3.2}$$

where $\epsilon(t)$, measured in people/time, represents the rate of importation of infections into the organization. A key challenge is that this function is not known in general. We assume proportionality to local case counts, and note that county-level casec counts are reported daily. Thus, be observable to decision makes and policy deciders. The continuous dependence on time means that we can test various scenarios (which gives more realism and flexibility) than the periodic forcing induced by the "exogenous shocks" considered by Paltiel et al. [2].

In the manuscript, we examine two data-driven scenarios for this forcing, but many possibilities can be incorporated into the model. The two extremes are meant to illustrate one of the key sources of uncertainty in any organization-based testing strategy — the number of infections entering the organization from outside.

- The first is a relatively flat profile which comes from the 7 day rolling average of the case count in Fayette County, Pennsylvania for the 100 days beginning March 26, 2020 as reported in the New York Times. We scale the case counts by population, which, in the

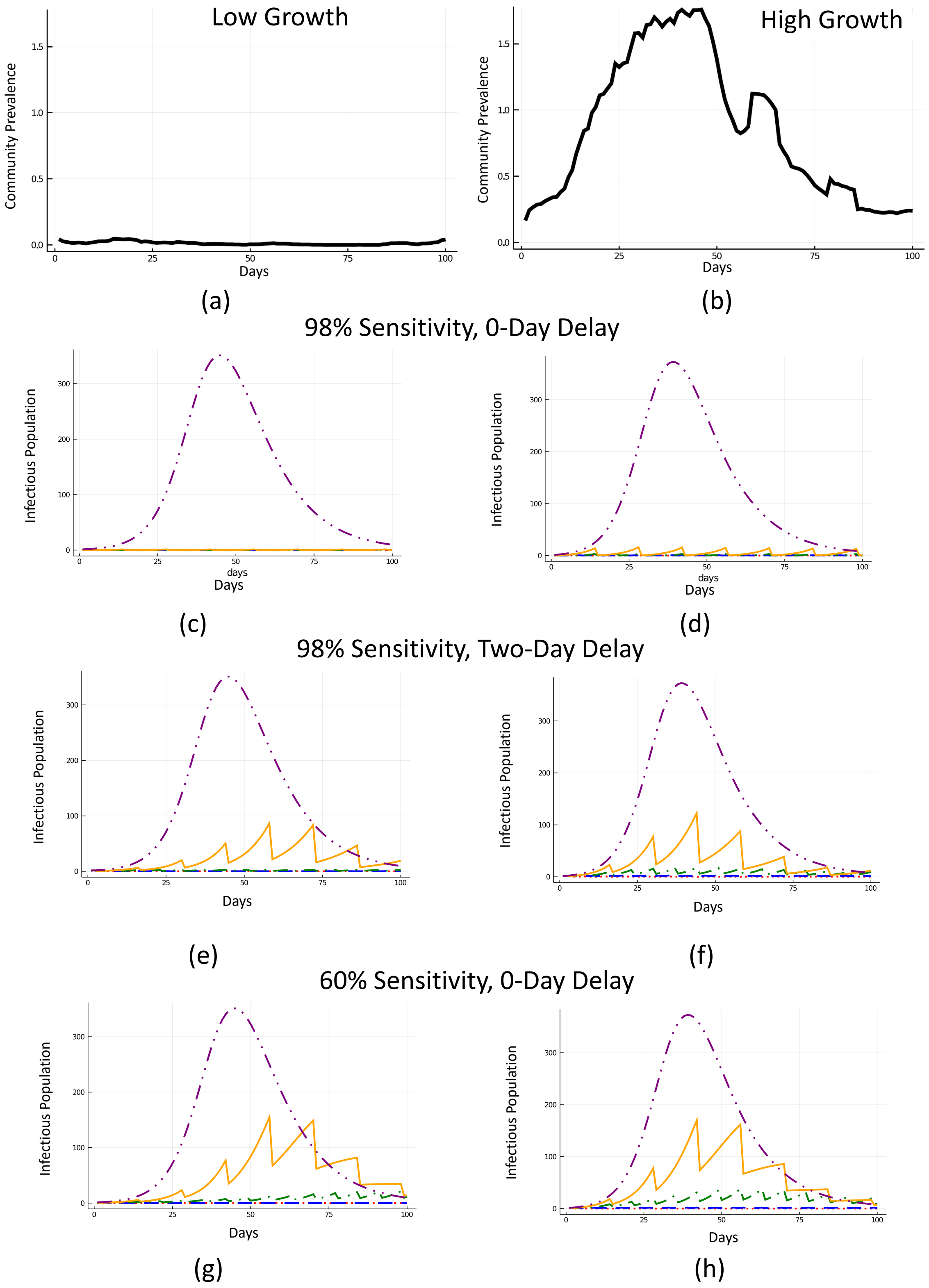


Figure 2. Reproduction of figure 2 in the manuscript without cropping.

manuscript we choose to be 1500. This low growth profile is reported as panel (a) in Figure 3.

- The second scenario used for high growth external community prevalence is the seven day rolling average of daily case counts in Miami-Dade County, Florida for the 100 days beginning June 16, 2020. This profile is shown in panel (b) of Figure 3.

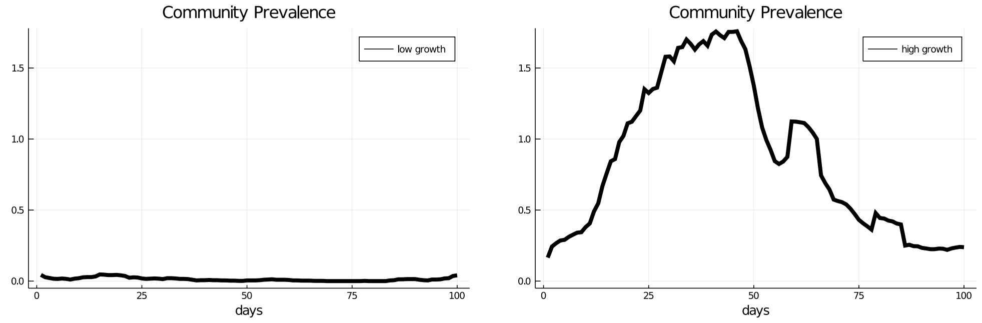


(a) (b)

FIGURE 3. Two examples of community prevalence. (a) Fayette County, PA. (b) Miami-Dade County, FL.

### 4. Core Julia code

The code representing heart of the dynamic model is reproduced below. Complete code, for creating all of the figures and creating the full table of experiments is available from the authors.

```
using DifferentialEquations
using Plots
using Printf
using Plots.PlotMeasures
using Statistics
using DataFrames
using CSV
using Interpolations
#read in profiles
profiles = DataFrame!(CSV.File("flat_growth.csv", types = Dict("fips" => String,
    "total_population"=>Float64)))

flat=Array(profiles[1,4:end])
growth=Array(profiles[2,4:end])

my_profile= "flat"#"growth"#


R0 = 0. # number of people recovered initially
R_0 = 2.5; #reproduction number
P = 1500;
gamma = 1/9.;
beta = R_0*gamma;
#1e-4 comes from the average number of new cases per person reported by NYT on
```

```
#9/23 in a representative sample of 511 counties
# *1/gamma since it takes that long to get out of infectious
I0 = 1e-4*P/gamma # number of people infected initially
T = 100 #number of days covered
r = 3.23e-3#discount rate for pooling
#https://www.medrxiv.org/content/10.1101/2020.04.19.20071639v2.full.pdf
#r = .4757 #saliva https://www.medrxiv.org/content/10.1101/2020.09.02.20183830v1

flat_itp=interpolate(P*flat,BSpline(Linear()))
flat_itp=scale(flat_itp,0:1:99)
flat_itp=extrapolate(flat_itp,Flat())
plot(flat_itp,label="low growth",title="Community Prevalence",xlabel="days",
    lw=5,ylims=(-.05,1.78),color=:black)
savefig("./figs/flat_profile.pdf")
g_itp=interpolate(P*growth,BSpline(Linear()))
g_itp=scale(g_itp,0:1:99)
growth_itp=extrapolate(g_itp,Flat())
plot(growth_itp,label="high growth",title="Community Prevalence",xlabel="days",
    lw=5,ylims=(-.05,1.78),color=:black)
savefig("./figs/growth_profile.pdf")
plot(flat_itp,label="low growth",lw=5,color=:blue, linestyle=:dash)
plot!(growth_itp,label="high growth",lw=5,color=:green, linestyle=:dashdot)
savefig("./figs/flat_and_growth_profile.pdf")


function sird(dq, q, alpha, t)
    gamma, beta, P, profile = alpha
    S = q[1]
    I = q[2]
    R = q[3]

    if profile=="flat"
        e_interp=interpolate(P*flat,BSpline(Linear()))
        e=scale(e_interp,0:1:99)
        epsilon=extrapolate(e,Flat())
    elseif profile=="growth"
        e_interp=interpolate(P*growth,BSpline(Linear()))
        e=scale(e_interp,0:1:99)
        epsilon=extrapolate(e,Flat())
    else
        println("ERROR: possible profiles are flat or growth")
    end

    dq[1] = -(beta * S) * I / P - epsilon(t)#S'
    dq[2] = (beta * S) * I / P - gamma * I + epsilon(t) #i'
    dq[3] = gamma * I #r'
end

function SolveSIRD(
```

```
    gamma::Float64,
    beta::Float64,
    I0::Float64,
    R0::Float64,
    P,
    tf::Int64,
    epsilon,
)
    S0 = P - I0 - R0
    ICS=[S0; I0; R0]

    prob = ODEProblem(sird, ICS, (0.0, tf),[gamma, beta, P, epsilon])

    sol = solve(prob, isoutofdomain=(u,p,t)->any(x->x<0,u))
    return sol
end

function find_SIR(I0,R0,r,m,epsilon,alpha_p0,tau::Int64, d::Int64, b::Float64,
        sp::Float64)
    #I0 number of initial infections
    #R0 number of initial removed
    #r discount rate for pooling samples
    #m number of samples pooled
    #epsilon people/day infected from the outside
    #alpha_p0 sensitivity of the test on 1 sample
    #d is the number of days delay between a test and results
    #b is the proportion of people who isolate upon positive results
    #sp is the specificity of the test

    S=Array{Float64}(undef, T+d+1)
    I=similar(S)
    R=similar(S)
    I_c = 0 #number of infections caught by testing
    fp = 0 #number of false positive tests
    PP = 0 #number of positive pools

    #first time step
    sol_n=SolveSIRD(gamma, beta, I0 ,R0, P, 1+d, epsilon)
    S[1:1+d] = sol_n(0:1:d)[1,:];
    I[1:1+d] = sol_n(0:1:d)[2,:];
    R[1:1+d] = sol_n(0:1:d)[3,:];

    #assumes prevalence in the pooled population = prevalence in the whole
        #population
    alpha_p=alpha_p0-r*(m-1)*(1-I[1]/P)
    #note that testing is done on day 1
    I0 = maximum([I[1+d]-b*alpha_p*I[1],0]);
    R0 = R[1+d]+b*alpha_p*I[1];
    I_c+=alpha_p*I[1] #number of infections caught
```

```
    fp +=(1-sp)*(P-I[1])
    prev=I[1]/P #prevalence
    pi_m=1-(sp+prev*(1-alpha_p-sp))^m
    PP+=ceil(P/m)*pi_m#+(1-pi_m))

    for n in 1:Int64(floor(T/tau))
        sol_n=SolveSIRD(gamma, beta, I0 ,R0, P, tau, epsilon)
        S[(n-1)*tau+d+1:n*tau+d+1] = sol_n(0:1:tau)[1,:];
        I[(n-1)*tau+d+1:n*tau+d+1] = sol_n(0:1:tau)[2,:];
        R[(n-1)*tau+d+1:n*tau+d+1] = sol_n(0:1:tau)[3,:];
        #assumes prevalence in the pooled population = prevalence in the whole
            #population
        alpha_p=alpha_p0-r*(m-1)*(1-I[n*tau+1]/P)

        I0 = maximum([I[n*tau+d+1]-b*alpha_p*I[n*tau+1],0]);
        R0 = R[n*tau+d+1]+b*alpha_p*I[n*tau+1];
        I_c+=alpha_p*I[n*tau+1] #number of infections caught
        fp +=(1-sp)*(P-I[n*tau+1])
        prev=I[n*tau+1]/P #prevalence
        pi_m=1-(sp+prev*(1-alpha_p-sp))^m
        PP+=ceil(P/m)*pi_m
    end
    if Int64(floor(T/tau)*tau) < T
        n=Int64(floor(T/tau)*tau)#last test day
        final_t_length = T-n
        alpha_p=alpha_p0-r*(m-1)*(1-I[n+1]/P)
        I0 = maximum([I[n+d+1]-b*alpha_p*I[n+1],0]);
        R0 = R[n+d+1]+b*alpha_p*I[n+1];
        I_c+=alpha_p*I[n+1] #number of infections caught
        #println(I_c)
        fp +=(1-sp)*(P-I[n+1])
        prev=I[n+1]/P #prevalence
        pi_m=1-(sp+prev*(1-alpha_p-sp))^m
        PP+=ceil(P/m)*pi_m

        sol_n=SolveSIRD(gamma, beta, I0 ,R0, P, final_t_length, epsilon)
        S[n+d+1:n+d+final_t_length] = sol_n(0:1:final_t_length-1)[1,:];
        I[n+d+1:n+d+final_t_length] = sol_n(0:1:final_t_length-1)[2,:];
        R[n+d+1:n+d+final_t_length] = sol_n(0:1:final_t_length-1)[3,:];
    end
    return S[1:T], I[1:T], R[1:T], I_c, fp, PP
end
#find_SIR(I0,R0,r,m,epsilon,alpha_p0,tau::Int64, d::Int64, b::Float64,sp)
S_t,I_t,R_t,I_c,fp,PP=find_SIR(I0,R0,r, 10, my_profile, .98,3, 0,1., .95)
C=125*ceil(P/10)*floor(T/3)+100*PP*10

#cost of doing nothing
@time sol_nt=SolveSIRD(gamma, beta, I0, R0, P, T,my_profile)
```

```
S_nt = sol_nt(0:1:T-1)[1,:];
I_nt = sol_nt(0:1:T-1)[2,:];
R_nt = sol_nt(0:1:T-1)[3,:];
```

### References


[1] Christopher R. Bilder. Group Testing for Identification. In *Wiley StatsRef: Statistics Reference Online*, pages 1–11. American Cancer Society, 2019. doi: 10.1002/9781118445112.stat08227.

[2] A. David Paltiel, Amy Zheng, and Rochelle P. Walensky. Assessment of SARS-CoV-2 Screening Strategies to Permit the Safe Reopening of College Campuses in the United States. *JAMA Network Open*, 3(7):e2016818–e2016818, July 2020.

[3] Smith, David RM, Duval, Audrey, Pouwels, Koen B, Guillemot, Didier, Fernandes, Jerome, Huynh, Bich-Tram, Temime, Laura, and Opatowski, Lulla. Optimizing COVID-19 surveillance in long-term care facilities: a modelling study. Preprint, August 2020.

[4] Idan Yelin, Noga Aharony, Einat Shaer Tamar, Amir Argoetti, Esther Messer, Dina Berenbaum, Einat Shafran, Areen Kuzli, Nagham Gandali, Omer Shkedi, Tamar Hashimshony, Yael Mandel-Gutfreund, Michael Halberthal, Yuval Geffen, Moran Szwarcwort-Cohen, and Roy Kishony. Evaluation of COVID-19 RT-qPCR Test in Multi sample Pools. *Clinical Infectious Diseases*, 2020.
